# Determinants of treatment success among drug-susceptible tuberculosis patients in Ghana: A prospective cohort study

**DOI:** 10.1101/2023.11.02.23298001

**Authors:** Richard Delali Agbeko Djochie, Berko Panyin Anto, Mercy Naa Aduele Opare-Addo

**Author notes:** Corresponding author, (BPA).

## Abstract

**Background:** Achieving optimal rates of success in tuberculosis treatment is vital not just for the individual patient’s well-being but also for preventing the rise of drug-resistant tuberculosis strains. Unfavourable treatment outcomes pose significant challenges for healthcare systems worldwide. Identifying the factors that determine treatment success is crucial for implementing effective interventions that enhance treatment outcomes and contribute to the eradication of the disease. As a result, this study aimed to assess tuberculosis treatment outcomes and identify factors associated with treatment success.

**Methods:** Patients diagnosed with active tuberculosis were closely monitored from the start of their treatment until its completion from January 2021 to June 2022. A data collection tool, developed using Redcap and aligned with the study objectives, was utilized to gather demographic information, and adverse reactions to antitubercular medicines and track the treatment outcomes of the participants. The participants’ quality of life was assessed using the Short-form 12 version 2 questionnaire at baseline, as well as at the end of the second and sixth months. Logistic regression was employed to evaluate the association between various participant characteristics and treatment success, with odds ratios used to quantify the strength of the associations.

**Results:** Among 378 participants, 77.3% had successful TB treatment, while 13.5% were lost to follow-up, 0.5% experienced treatment failure, and 8.7% died. Factors influencing treatment success included initial body weight, weight gain during treatment, HIV status, drug adverse reactions, and mental well-being at the beginning of treatment. Multivariate analysis showed that gaining at least 3kg during treatment and having no risk of depression at the beginning significantly increased the likelihood of successful treatment.

**Conclusion:** Patients with tuberculosis who experience compromised physical and mental health-related quality of life, encounter adverse reactions to antitubercular drugs and have concurrent HIV infection should receive close monitoring and personalized interventions to improve their chances of treatment success.

## Introduction

Tuberculosis (TB) remains a pressing global health concern, affecting more than 10 million people each year [1]. Despite being a preventable and treatable infection, TB ranks as the second deadliest disease worldwide, only behind COVID-19 with 1.5 million fatalities in 2021 alone. The majority of TB cases occur in low- and middle-income countries within the World Health Organisation’s (WHO) regions of South-East Asia, Africa, and the Western Pacific [1]. In 2021, Ghana reported 136 tuberculosis cases per 100,000 individuals, resulting in approximately 12,000 deaths attributed to TB only and 3,700 TB/HIV coinfection deaths [2]. The impact of the COVID-19 pandemic on healthcare services may have exacerbated this situation [1].

Tuberculosis treatment success rates exhibit significant variations across different countries and regions. As of 2020, the global TB treatment success rate stood at 85%, short of the End TB Strategy target of 90% by 2025 [1]. In the United States, it was reported at 74%, while the African Region averaged at 79% [1,3]. Adverse treatment outcomes for drug-susceptible TB cases pose a significant challenge to healthcare systems worldwide. Achieving optimal treatment success rates is not only essential for individual patient well-being but also for preventing the emergence of drug-resistant TB strains. Despite free TB treatment accessibility in Ghana, the mortality rate associated with this disease remains a concern. Since 2012, Ghana’s TB treatment success rate has plateaued at 84-85% [4,5], falling short of WHO’s recommended target of 90% [6]. Studies in Ghana, primarily retrospective in design, report varying treatment success rates across regions: 82.5% in the Volta region, 90.2% in the Central region, and 90.7% in the Greater Accra region [7,8]. Prioritizing strategies to enhance treatment success is vital for the National Tuberculosis Control Program (NTP) to reduce re-treatment needs, save costs, and lower morbidity and mortality rates.

In lower- to medium-income countries, studies have identified factors contributing to unfavourable TB treatment outcomes, including limited access to health logistics, nonadherence to treatment, HIV coinfection, alcohol use, and drug abuse [9–11]. Other authors have also highlighted a premature sense of recovery before completing the treatment, the absence of social support, persistent stigma, and limited knowledge about TB as factors associated with adverse treatment outcomes [12,13]. A systematic review found socioeconomic elements such as inadequate housing (characterized by overcrowding and poor ventilation), malnutrition, and alcohol abuse as negative influences on TB treatment outcomes [14].

Tuberculosis treatment success is influenced by a complex interplay of sociodemographic and clinical factors including low income, substance abuse, gender disparities, education level, unemployment, advanced age, membership in migratory communities, and the stigma surrounding TB, which collectively shape the outcomes of TB treatment programmes [14–16]. Evaluating TB treatment outcomes and identifying the root causes is essential for health authorities and policymakers.

While numerous studies have delved into the factors influencing low treatment success rates in Ghana, it’s important to note that much work remains to be done in this field. Although some contributing factors to successful treatment outcomes, such as age and pretreatment weight, have been identified, there is still a critical need to expand the scope of research in order to gain a more comprehensive understanding of these complex dynamics [12,13,17]. Most of these studies, predominantly retrospective in design, have yet to fully consider the impact of health-related quality of life and the occurrence of adverse reactions to anti-TB drugs on treatment outcomes. Therefore, there is a need for further research efforts to explore these factors in greater depth, ultimately paving the way for more effective interventions to improve TB treatment success rates in Ghana and similar settings. Consequently, this study aimed to assess treatment outcomes and identify factors associated with success in patients with drug-susceptible TB taking into consideration the impact of anti-TB adverse drug reactions and health-related quality of life. The findings will contribute to the body of knowledge in TB management and provide evidence for health managers and policymakers to develop tailored interventions for specific demographics. These interventions aim to tackle the less-than-optimal rates of treatment success and ultimately reduce the burden of TB within the country and similar global settings.

## Methods

### Study design and participants

This study was conducted in the Eastern and Ashanti Regions of Ghana, specifically at eight tuberculosis treatment centres. These centres were chosen based on their consistent reporting of the highest number of TB cases annually in their respective regions over the past five years. The study sites consisted of three primary care hospitals and one regional referral hospital. The study population included newly diagnosed TB patients who were eligible for drug treatment and ready to initiate TB treatment. However, only patients who were 18 years of age or older, had access to a mobile phone and provided their consent by signing a consent form were included. Conversely, individuals with psychiatric conditions, liver or kidney impairment were excluded from the study.

To ensure sufficient statistical power with an 80% power analysis, a two-tailed test, an alpha level of 0.05, and a medium effect size of 0.5, a minimum of 128 participants was required for this study. However, considering potential non-responses and ensuring a diverse representation of tuberculosis patients, 378 respondents were recruited.

### Data collection and analysis

The data collection instrument utilized in this study was developed using REDCap, aligning with the study objectives. To ensure its reliability, the instrument underwent validation and was pretested in a hospital that was not part of the main study. The feedback received from the pretest was utilized to make necessary adjustments to the instrument.

Participant recruitment and data collection were conducted from 10^th^ January 2021 to 15^th^ April 2022. Trained research assistants called each participant, contacting them at least once a week to inquire about any adverse effects resulting from their TB treatment. Additionally, any adverse drug reactions (ADRs) recorded on the participants’ TB treatment cards at the clinic were extracted. A list of side effects associated with antitubercular drugs, documented in the literature, was used to screen the participants. The Naranjo ADR Probability Scale was employed to determine the causal relationship between the reported ADRs and the antitubercular drugs. Confirmation of a causal relationship was established in scenarios where a patient experienced the same reaction after re-administration of the drug, when the reaction only occurred following drug administration, when the reaction subsided or ceased after the administration of an antidote, and when the patient was not taking any alternative medication known to cause the same reaction [18]. Only ADRs classified as Certain (a score of ≥9), Probable (a score of 5-8), or Possible (a score of 1-4) were included in the analysis. The severity of antitubercular drug-related adverse reactions (ADRs) in participants was assessed using Hartwig’s ADR severity scale and categorized as severe (levels 5-7) if they led to death, in-patient hospitalization, or prolonged hospital stay; moderate (levels 3-4) if they required withholding or discontinuation of anti-TB treatment and/or an antidote; and mild (levels 1-2) if they necessitated suspending anti-TB drugs without an antidote [19,20]. For analysis purposes, moderate-to-severe ADRs were grouped as major ADRs, while mild ADRs were considered minor ADRs.

The Short-form 12 version 2 questionnaire (SF-12v2) was employed in this study to assess the participants’ quality of life at baseline, as well as at the end of the second and sixth months. The SF-12v2 survey consists of 12 items that measure various aspects of functioning and well-being from the patient’s perspective. This questionnaire is a validated and widely accepted tool for evaluating the quality of life in individuals aged 18 and above, particularly in the context of chronic illnesses [21,22]. The validity of the SF-12v2 as a health-related quality-of-life evaluation instrument has also been confirmed among Ghanaian adults [23]. For comparative analysis, the physical component summary (PCS) and mental component summary (MCS) scores were utilized in the current study [22]. A higher PCS or MCS score indicates a better quality of life. A minimum clinically important difference (MCID) in quality of life at the end of treatment was defined as a 3-point increase in PCS or MCS scores while participants with an MCS score below 42 were classified as experiencing depression [22].

### Statistical analysis

Data cleansing was executed with Microsoft Excel 2016, and subsequently, data analysis was conducted using IBM SPSS Statistics (version 21). The findings were presented in the form of frequencies, percentages, and means with standard deviations. Chi-squared or Fisher’s exact test, was applied to identify socio-demographic and clinical factors linked to treatment success. Logistic regression analysis, incorporating both crude and adjusted odds ratios, was utilised to gauge the strength of any observed associations between patient characteristics and treatment success. The threshold for statistical significance was established at less than 0.05.

### Ethical considerations

The study was conducted following ethical guidelines and was approved by two committees. The Ghana Health Service Ethics Review Committee granted approval with the number GHS-ERC 007/11/20 on January 2, 2021, and the Committee on Human Research, Publications, and Ethics of Kwame Nkrumah University of Science and Technology approved it with the number CHRPE/AP/171/21 on April 27, 2021.

Throughout the study, ethical principles were upheld. These principles included ensuring voluntary participation, beneficence, nonmaleficence, privacy, and confidentiality. Participants were fully informed and willingly chose to participate. They also provided written informed consent. Importantly, participants had the right to withdraw from the study at any time, and they were not required to provide a reason for doing so.

## Results

### Sociodemographic characteristics of participants

Among the 378 participants, the majority were male (67.2%), married (44.2%), and employed (75%), with a mean age of 45.3 (±15.1) years. As shown in Table 1, 12.9% were illiterate, and a small proportion of participants had extrapulmonary TB (6.1%), were smokers (6.4%), consumed alcohol (25.4%), or were HIV-positive (25.7%).

**Table 1.** Sociodemographic and clinical characteristics of study participants.

| **Characteristics** | **Frequency**<br>***n*** | **Percentage of**<br>**total** |
| --- | --- | --- |
| **Age (years)** |  |  |
| 18 – 25 | 26 | 6.9 |
| 26 – 35 | 84 | 22.2 |
| 36 – 45 | 90 | 23.8 |
| 46 – 55 | 90 | 23.8 |
| 56 – 65 | 49 | 13.0 |
| ≥66 | 39 | 10.3 |
| **Gender** |  |  |
| Male | 254 | 67.2 |
| Female | 124 | 32.8 |
| **Marital status** |  |  |
| Married | 167 | 44.2 |
| Single | 127 | 33.6 |
| Divorced | 41 | 10.9 |
| Widowed | 43 | 11.3 |
| **Employment status** |  |  |
| Employed | 289 | 76.5 |
| Unemployed | 89 | 23.5 |
| **Educational level** |  |  |
| Basic | 230 | 60.9 |
| Secondary | 70 | 18.5 |
| Tertiary | 29 | 7.7 |
| None | 49 | 12.9 |
| **Type of TB** |  |  |
| Pulmonary TB | 350 | 92.6 |
| Extra-pulmonary TB | 23 | 6.1 |
| Both | 5 | 1.3 |
| **Smoking status** |  |  |
| Smoker | 24 | 6.4 |
| Nonsmoker | 354 | 93.6 |
| **Alcohol use** |  |  |
| Yes | 96 | 25.4 |
| No | 282 | 74.6 |
| **HIV status** |  |  |
| HIV positive | 97 | 25.7 |
| HIV negative | 275 | 72.8 |
| HIV status unknown | 6 | 1.5 |
| **Baseline HRQOL** |  |  |
| Depression | 97 | 25.7 |
| Physical impairment | 298 | 78.8 |
| **ADR Status** |  |  |
| No ADR | 198 | 52.4 |
| Minor ADR | 68 | 18.0 |
| Major ADR | 112 | 29.6 |

TB = Tuberculosis, HIV = Human immune-deficiency virus, ADR = Adverse drug reaction, HRQOL = health-related quality of life

### Treatment outcomes of study participants

The distribution of tuberculosis treatment outcomes among the study participants was as follows: 44.2% were cured, 33.1% completed their therapy, 13.5% were lost to follow-up, 8.7% died, and 0.5% were classified as treatment failures. Therefore, the overall treatment success rate in this study was 77.3%.

As illustrated in Table 2, gender did not significantly affect treatment success, with 78.7% of males and 74.2% of females achieving successful treatment (p = 0.322). However, a notable difference was observed based on participants’ baseline weight, where those with a weight of at least 53kg had a significantly higher success rate (83.6%) compared to those with less weight (72.6%) (p = 0.011). Furthermore, the amount of weight gained during treatment had a substantial influence on treatment success. Participants who gained a minimum of 3kg during treatment demonstrated a remarkable success rate of 90.9%, while those who gained less than 3kg had a success rate of 65.5% (p < 0.001). Other factors, such as alcohol use, tobacco smoking, and the initiation of Highly Active Antiretroviral Treatment (HAART) before tuberculosis, did not show statistically significant associations with treatment success. However, participants with HIV-negative status exhibited a significantly higher success rate (81.8%) compared to those with HIV-positive status (64.9%) (p = 0.002). Additionally, the type of HIV infection (HIV-1, HIV-2, or both) had a significant impact on treatment success, with HIV-2-infected participants demonstrating the lowest success rate (57.1%) compared to the other groups (p = 0.011). Regarding the site of TB infection, those with extrapulmonary TB achieved a 100% success rate, while participants with pulmonary TB had a success rate of 75.4% (p = 0.003).

**Table 2.** Association between patient characteristics and treatment success.

| Parameter | Number of participants | % of patients with treatment success | p-value |
| --- | --- | --- | --- |
| **Gender** |  |  |  |
| Male | 254 | 78.7 | 0.322 |
| Female | 124 | 74.2 |  |
| **Weight at baseline** |  |  |  |
| < 53kg | 219 | 72.6 | 0.011 |
| ≥ 53kg | 159 | 83.6 |  |
| **Total weight gained** |  |  |  |
| < 3kg | 174 | 65.5 | <0.001 |
| ≥ 3kg | 170 | 90.9 |  |
| **Alcohol use** |  |  |  |
| Yes | 96 | 72.9 | 0.241 |
| No | 282 | 78.7 |  |
| **Tobacco smoking** |  |  |  |
| Yes | 24 | 83.3 | 0.462 |
| No | 354 | 76.8 |  |
| **HIV status** |  |  |  |
| HIV-positive | 97 | 64.9 | 0.002 |
| HIV-negative | 275 | 81.8 |  |
| HIV status unknown | 6 | 66.7 |  |
| **HIV Type** |  |  |  |
| HIV-1 | 83 | 69.9 | 0.011 |
| HIV-2 | 7 | 57.1 |  |
| Both HIV-1 & 2 | 7 | 14.3 |  |
| **Site of TB infection** |  |  |  |
| Pulmonary | 350 | 75.4 | 0.003 |
| Extra-pulmonary | 28 | 100.0 |  |
| **HAART started before TB** |  |  |  |
| Yes | 84 | 72.0 | 0.133 |
| No | 13 | 57.4 |  |
| **ADR status** |  |  |  |
| No ADR | 198 | 77.3 | 0.991 |
| At least one ADR | 180 | 77.2 |  |
| **Physical HRQOL at baseline** |  |  |  |
| Good physical health | 76 | 88.8 | 0.006 |
| Poor physical health | 302 | 74.2 |  |
| **Mental HRQOL at baseline** |  |  |  |
| Good mental health | 281 | 80.2 | 0.026 |
| Poor mental health | 97 | 69.1 |  |

WHO=World Health Organization; MCS=mean component summary; PCS=physical component summary; HIV=human immunodeficiency virus; HAART=highly active antiretroviral treatment; TB=tuberculosis; ADR = adverse drug reaction.

The presence or absence of adverse reactions to antitubercular drugs didn’t make a significant difference in the success rates of treatment. The success rates were quite similar at 77.3% for those with adverse reactions and 77.2% for those without (p = 0.991). Additionally, the severity of these adverse reactions also did not affect the treatment outcomes.

However, when looking at participants who experienced minor and major adverse reactions separately, their success rates were 80.9% and 75.0%, respectively (p = 0.362). So, the severity of the adverse reactions didn’t seem to impact success significantly.

On the other hand, the initial physical health of participants did have a notable impact on the treatment’s success. Those in good physical health had a significantly higher success rate of 88.8%, compared to those in poor physical health, who had a success rate of 74.2% (p = 0.006). Similarly, the initial mental health of participants also played a role, with those in good mental health achieving a higher success rate of 80.2%, while those in poor mental health had a lower success rate of 69.1% (p = 0.026).

### Determinants of treatment success

The robustness of the relationships between patient characteristics and the success of treatment, as depicted in Table 3, reveals significant findings. Participants with a baseline weight above 53 kg exhibited 1.9 times higher odds of achieving treatment success compared to those with a weight of 53 kg or less, with an odds ratio (OR) of 1.9 (95% CI: 1.2 – 3.2, p = 0.012). Additionally, individuals who gained at least 3 kg during the course of treatment demonstrated significantly increased odds of treatment success, with an OR of 4.5 (95% CI: 2.2 – 9.0, p < 0.001).

**Table 3.** The effect of participants’ characteristics on the odds of having treatment success.

| Patient characteristics | Treatment success |  |  |
| --- | --- | --- | --- |
|  | OR | 95% CI | P-value |
| **Weight at baseline** |  |  |  |
| ≤ 53 kg | 1 |  |  |
| >53 kg | 1.9 | 1.2 – 3.2 | 0.012 |
| **Total weight gained during treatment** |  |  |  |
| <3 kg | 1 |  |  |
| ≥3 kg | 4.5 | 2.2 – 9.0 | <0.001 |
| **HIV status** |  |  |  |
| HIV-positive | 1 |  |  |
| HIV-negative | 2.4 | 1.4 – 4.1 | <0.001 |
| **Type of HIV** |  |  |  |
| HIV-2 | 1 |  |  |
| HIV-1 | 4.2 | 1.3 – 3.7 | 0.019 |
| **Physical HRQOL at baseline** |  |  |  |
| Poor physical health | 1 |  |  |
| Good physical health | 2.5 | 1.2 – 5.3 | 0.014 |
| **Mental HRQOL at baseline** |  |  |  |
| Poor mental health | 1 |  |  |
| Good mental health | 1.8 | 1.1 – 3.0 | 0.027 |
| **Gastrointestinal ADR** |  |  |  |
| Yes | 1 |  |  |
| No | 1.8 | 1.1 – 3.0 | 0.019 |
| **Hepatobiliary system ADR** |  |  |  |
| Yes | 1 |  |  |
| No | 21.8 | 2.6 – 183.0 | 0.005 |

ADR = adverse drug reaction; HRQOL = health-related quality of life; HIV = human immunodeficiency virus

HIV status played a crucial role in treatment outcomes. HIV-negative participants had more than twice the odds of achieving treatment success compared to their HIV-positive counterparts, with an OR of 2.4 (95% CI: 1.4 – 4.1, p < 0.001). Furthermore, participants with HIV-1 infection showed over four times the odds of success compared to those with HIV-2, with an OR of 4.2 (95% CI: 1.3 – 3.7, p = 0.019).

The physical and mental HRQOL at the beginning of treatment significantly influenced outcomes. Participants in good physical health had three times higher odds of treatment success compared to those in poor physical health, with an OR of 2.5 (95% CI: 1.2 – 5.3, p = 0.014). Similarly, individuals in good mental health exhibited nearly twice the odds of success compared to those in poor mental health, with an OR of 1.8 (95% CI: 1.1 – 3.0, p = 0.027).

Furthermore, antitubercular ADRs had varying impacts on treatment success. While the severity of ADRs did not significantly affect treatment outcomes, participants who did not experience gastrointestinal ADRs had twice the odds of treatment success (OR = 1.8, 95% CI: 1.1 – 3.0, p = 0.019), and those without hepatobiliary system ADRs had a strikingly higher likelihood of treatment success, with odds almost 22 times greater than those with such ADRs (OR = 21.8, 95% CI: 2.6 – 183.0, p = 0.005).

A multivariate analysis considering all statistically significant factors related to treatment success indicated that gaining at least 3kg of body weight at the end of treatment (p = 0.004) and not being at risk of depression at the beginning of treatment (p = 0.016) remained significantly associated with a successful treatment outcome.

While this study observed that a higher proportion of males, non-smokers, non-drinkers, and patients receiving Highly Active Antiretroviral Treatment (HAART) tended to have improved treatment outcomes, it’s important to note that these distinctions did not attain statistical significance.

## Discussion

The TB treatment success rate in this study stood at 77.3%, comprising 44.2% of participants who were cured and 33.1% who completed their treatment. This rate fell below both the national average of 84% [6] and the WHO’s target of 90% [8]. It also trailed behind the success rates of 81.8%, 81.0% and 90.2% reported in studies conducted in Somalia [24], India [26], and another study within the Central region of Ghana [7]. Nonetheless, it exceeded the success rates reported in certain Ethiopian studies (71.4% and 60.1%) [25,27] as well as the 68.5% reported by a ten-year retrospective study conducted in the Ashanti Region of Ghana [17].

Among the participants, 22.8% experienced adverse treatment outcomes, including 13.5% who were lost to follow-up, 8.7% who died, and 0.5% who had treatment failure. The loss to follow-up rate in this study was similar to the 6.2% observed in Somalia [24] and the 8% reported in Ethiopia [25]. However, it was lower than the 21.3% found in another Ethiopian study [27] and the 12.5% in an Indian study [26] but higher than the 0.9% reported in another Ghanaian study [7]. Defaulting on TB treatment not only worsens symptoms to the point of requiring hospitalization but also poses a public health risk by potentially spreading drug-resistant strains of Mycobacterium. Therefore, it is crucial to provide comprehensive pre-treatment adherence counselling and continue reinforcing it during clinic visits [28].

Among the 33 participants who died during TB treatment, 63.6% (n=21) were HIV-positive. The TB death rate (8.7%) observed in this study was lower than the 10% target set by the TB strategic plan [29], 17.7% reported in Ethiopia [27], and 10.1% in another Ethiopian study [30]. However, it was higher than the 4.17% in a Pakistani study [31] and the 2% reported in India [26]. Tuberculosis infection accelerates the progression of acquired immune deficiency syndrome (AIDS) in HIV-positive individuals [32], making them more vulnerable to opportunistic infections that require additional treatment. In contrast, tuberculosis patients without HIV infection generally achieve better treatment outcomes [10,24,27,30,33]. This is due to the detrimental effect of HIV infection on CD4 cell counts and overall immunity, which impairs successful treatment. The higher pill burden and potential drug interactions associated with HIV treatment contribute to increased nonadherence rates and lower rates of treatment success.

Importantly, the type of HIV infection influences treatment outcomes, with HIV-1 patients having more than double the odds of achieving successful treatment outcomes compared to those with HIV-2 infection. This finding aligns with a prospective study conducted in Guinea Bissau, which found that HIV-infected patients showed greater clinical severity of tuberculosis [32]. It underscores the significance of promptly diagnosing and treating HIV in tuberculosis patients, as well as offering continuous adherence support and psychological counselling during treatment for individuals living with HIV, to ensure treatment success.

Unlike certain studies conducted in the African region [25,27], this study reveals a notable positive correlation between extrapulmonary TB and treatment success. In our study, all participants with extrapulmonary TB achieved successful treatment outcomes, aligning with findings from analogous studies conducted in Bhutan and Ethiopia [34,35]. Further research is needed to comprehensively understand the relationship between the type of TB infection and treatment success.

The importance of increasing body weight in TB patients through proper nutrition to improve their physical health and treatment outcomes has been widely recognized [36–39]. This study confirms that patients who had a baseline weight of over 53kg or gained at least 3kg of body weight during treatment achieved successful outcomes, consistent with findings from Peru and Ethiopia [39,40]. The positive impact of weight gain on treatment outcomes can be attributed to improved nutrition, which strengthens the immune system and supports the healing process. Therefore, offering nutritional support to vulnerable TB patients, such as those experiencing homelessness or living in impoverished households, could significantly enhance their chances of treatment success.

Participants who did not have physical impairment at the beginning of treatment had nearly three times higher odds of achieving a successful treatment outcome compared to those with physical impairment. Similarly, patients without a risk of depression at baseline had almost twice the odds of treatment success. It is crucial for healthcare workers dealing with TB to identify such patients and provide them with adequate counselling to address the psychosocial and emotional challenges associated with the disease. Additionally, effectively managing comorbidities that could cause physical impairment can improve patients’ physical strength and increase their likelihood of treatment success. Implementing interventions to eliminate stigma within communities is also highly recommended.

We found no significant association between the rate of treatment success among participants who did not experience any anti-TB drug reactions and those who had at least one ADR. Moreover, the difference in treatment success rate among participants who had minor ADRs and those who had major ADRs was not statistically significant. This suggests that the severity of adverse drug reactions did not appear to significantly influence treatment outcomes in this study. However, it was observed that participants who did not develop any gastrointestinal adverse drug reactions had twice the odds of treatment success compared to those who did. Similarly, participants who did not experience any hepatobiliary system adverse drug reactions had 22 times the odds of treatment success compared to those who did. These findings align with two prospective studies conducted in India and China, which also reported that the absence of antitubercular adverse drug reactions significantly correlated with treatment success [41,42]. To fully understand the role of gastrointestinal and hepatobiliary ADRs in determining TB treatment success, further research is required.

## Conclusion

This study highlights several key factors that significantly influence treatment outcomes in drug-susceptible tuberculosis cases: including baseline weight, weight gain during treatment, HIV status, mental and physical health, and the presence of gastrointestinal and hepatobiliary system adverse drug reactions. Recognizing and addressing these determinants can guide healthcare professionals in tailoring interventions to enhance treatment success rates, ultimately contributing to the battle against tuberculosis.

## Data Availability

All data sets associated with this manuscript are available at data.mendeley.com

www.data.mendeley.com

## Acknowledgement

The authors are grateful to the managers of the hospitals in the study sites and all TB patients who willingly participated in the study.

